# Dissemination of OXA-23 carbapenemase-producing *Proteus mirabilis* and *Escherichia coli* is driven by transposon-carrying lineages in the UK

**DOI:** 10.1101/2025.03.14.25323970

**Authors:** Roxana Zamudio, Karen Osman, Rachel Pike, Aiysha Chaudhry, Danièle Meunier, Nicole Stoesser, Rebecca Stretch, Jane F Turton, David Williams, Katie L Hopkins

## Abstract

Carbapenem-resistant Enterobacterales are a significant threat to global public health. Here, we characterize *bla*_OXA-23_-positive *Proteus mirabilis* (n=8) and *Escherichia coli* (n=3) isolates from human clinical samples collected between 2021–2024 in the UK. Whole genome sequencing (WGS) was used to generate data, and a core gene SNP-based phylogenetic tree was constructed to assess the genomic relatedness among the isolates. To provide an international context, we included publicly available genomes. Short-read mapping to a reference genome enabled reconstruction of the genomic neighborhood around *bla*_OXA-23_. Minimum inhibitory concentration (MIC) determination was performed using broth microdilution and results interpreted using EUCAST guidelines. UK *P. mirabilis* isolates belonged to ST142 and were closely related (2–13 SNPs) to French isolates from 2017-2019. *E. coli* ST38 isolates harboured *bla*_OXA-23_ and showed high genetic relatedness (5–9 SNPs) among themselves. In *P. mirabilis*, *bla*_OXA-23_ was associated with transposon Tn*6703*, while *E. coli* harboured a novel composite transposon, designated Tn*7816*, bordered by two copies of IS*15DIV* and with three copies of *bla*_OXA-23_. *bla*_OXA-23_ was integrated into the chromosome in all isolates. All isolates were resistant to amoxicillin/clavulanic acid (>32 mg/L) and with meropenem MICs above the EUCAST screening cut-off (0.5–1 mg/L). In conclusion, UK *bla*_OXA-23_-positive *P. mirabilis* isolates belong to the same clonal lineage (ST142) previously reported in Belgium, Germany, Switzerland and France, suggesting introduction of this lineage into the UK. This is the first report of an *E. coli* ST38 lineage with chromosomally-encoded *bla*_OXA-23_ located within a novel transposon Tn*7816*.

WGS plays an important role in identifying the mechanism(s) of transmission of emerging carbapenemase genes.

**IMPACT STATEMENT:** Most diagnostic assays primarily focus on detection of the ‘big 5’ carbapenemase gene families (KPC, OXA-48-like, NDM, VIM and IMP), which are globally dominant in Enterobacterales. OXA-23-like carbapenemase genes are predominantly identified in *Acinetobacter baumannii* meaning that their presence in Enterobacterales is likely underestimated. In this study, we report the identification of *bla*_OXA-23_ in *Proteus mirabilis* and *Escherichia coli* isolated from UK clinical samples following incorporation of *bla*_OXA-23-like_ as a target in the multiplex-PCR assay used to screen all Gram-negative bacteria referred to the UK’s national reference laboratory for investigation of carbapenem resistance. To enhance our understanding of the genomic epidemiology of *bla*_OXA-23_ we utilized short-read sequencing to characterize all isolates, and long-read assembly polished with short-read to determine the genomic context of *bla*_OXA-23_ in *E. coli*. WGS analysis provided invaluable insights into the genomic relatedness among the isolates, identifying that *bla*_OXA-23_-positive *P. mirabilis* isolates were closely related to those previously reported in Europe and uncovered a novel transposon associated with *bla*_OXA-23_ in *E. coli*. In addition, *bla*_OXA-23_ was chromosomally located in all isolates, with the potential for stable vertical inheritance. The results of this study highlight the need to further characterize Enterobacterales isolates suspected of carbapenemase production but negative for the ‘big 5’ carbapenemase gene families and further demonstrates the role of WGS in characterizing bacterial strains and mobile genetic elements associated with the emergence and transmission of antimicrobial resistance mechanisms.

**DATA SUMMARY:** Illumina short-read, contigs and MIC (if available) data for *P. mirabilis* (n=8) and *E. coli* (n=3) *bla*_OXA-23_-positive isolates from the UK are available in the ENA database under Bioproject PRJEB80458. The Illumina polished version of the long-read-only (Nanopore) assembly for 1697008 (ES1) *E. coli* isolate is accessible under the assembly accession number GCA_964341145. The accession number for each genome and the metadata generated in this study are provided in the Table 1.

## INTRODUCTION

Carbapenem-resistant Enterobacterales were designated as critical priority pathogens by the World Health Organization (WHO) because of their limited treatment options and widespread prevalence^1^. Carbapenem resistance in Enterobacterales is primarily driven by the acquisition of one or more of the ‘big 5’ carbapenemase genes, primarily β-lactamases of Ambler class A (*bla*_KPC_), class B (*bla*_NDM_, *bla*_VIM_, *bla*_IMP_) and class D (*bla*_OXA-48-like_)^2^. In contrast, *bla*_OXA-23_ is commonly present in *Acinetobacter baumannii* and has been disseminated globally^3^, but has only been occasionally reported in Enterobacterales. Chromosomally-encoded *bla*_OXA-23_ in *P. mirabilis* was first detected in 1996 in France and since then, isolates have been reported in various European countries in clinical^4–10^ and animal samples^5^. In contrast, reports of *bla*_OXA-23_ in *E. coli* are very rare, with the first clinical case identified in Singapore^11^, and 14 subsequent cases documented in India between 2013-2014^12^. Since 2020, the UK’s national reference laboratory has screened all Gram-negatives submitted for investigation of carbapenem resistance with a multiplex PCR including *bla*_OXA-23-like_, *bla*_OXA-40-like,_ *bla*_OXA-51-like_ and *bla*_OXA-58-like_ families. Here we describe the genomic characterization of the first UK *P. mirabilis* and *E. coli* isolates harbouring *bla*_OXA-23._

## MATERIALS AND METHODS

### Bacterial isolates and antimicrobial susceptibility testing

Isolates were referred to the UK Health Security Agency (UKHSA) Antimicrobial Resistance and Healthcare Associated Infections (AMRHAI) Reference Unit for investigation of carbapenem resistance following recovery from clinical human specimens (urine, rectal swab and wound) collected in the UK between 2021 and 2024. Antimicrobial susceptibility testing was performed using broth microdilution against AMRHAI’s standard antibiotic panel and results interpreted according to the European Committee on Antimicrobial Susceptibility Testing (EUCAST) clinical breakpoints version 14.0^13^.

### Whole genome sequence and genome collection

Paired-end short-read sequencing was performed using Illumina technology. The methods for DNA extraction and WGS were previously described^14^; briefly, DNA was extracted from RNAse-treated lysates using a QIAsymphony DSP DNA Midi kit (Qiagen, Hilden, Germany) and sequenced on a Hiseq 2500 instrument (Illumina, San Diego, CA, USA) using the standard 2×101 bp sequencing protocol. Additionally, a complete chromosome sequence determined by MinION sequencing was available for the *bla*_OXA-23_-positive 1697008 *E. coli* isolate (with the alternative name of ES1; GenBank: CP133856.1). Long-read sequencing was performed on a minION Mk1C device (Oxford Nanopore Technologies, Oxford, United Kingdom) using an R10.4.1 flow cell following library preparation using the rapid barcoding kit SQK-RBK114.24. Basecalling was performed using the high-accuracy model.

*bla*_OXA-23_-positive *P. mirabilis* genomes (n=56) from previous studies were included for international context (Supplementary Table S1). Draft genomes were available for most isolates: a subset of the French isolates had accessible Illumina short-reads, and a complete chromosome sequence was available for one isolate from France (named as VAC; GenBank: CP042907.1). Furthermore, 32 *bla*_OXA-23_-negative *E. coli* ST38 genomes from a previous UK study^15^, with Illumina short-read availability, were included for comparative genome analysis with the *bla*_OXA-23_-positive *E. coli* ST38 (Supplementary Table S1). More details are available in the supplementary methods (see Supplementary Material).

### Assembly, typing and screening of AMR determinants

Illumina reads were processed with Trimmomatic v0.39^16^ (default parameters) to eliminate adapters and poor-quality bases. Subsequently, the trimmed reads were utilized to assemble draft genomes using SPAdes v3.11.1^17^ with default parameters and the --careful option. Assemblies were evaluated using Quast v5.2.0^18^ for summary statistics, and quality was also assessed with CheckM v1.2.2^19^, where any genomes with less than 95% completeness and/or assembly contamination exceeding 2% were excluded. The long-read-only assembly obtained using Flye v2.9.1-b1780 and Medaka v1.7.2 for the 1697008 isolate (also known as ES1; Genbank CP133856.1) has been further polished using Illumina reads and Pilon v1.24^20^ to correct a small number of mismatches (31 SNPs and 40 indels, which represent 0.001% of the sequence) in the assembly. This Illumina polished version of the complete chromosome for the 1697008 isolate is available under assembly accession number GCA_964341145. This Illumina polished version was used in subsequent analysis. AMR genes and point mutations were identified in the assemblies using AMRFinderPlus v3.12.8^21^ with database version 2024-01-31.1^21^. Multi-locus sequence typing (MLST) schemes for *Proteus* spp. and *E. coli* were utilized to determine the sequence type (ST) of each isolate through the pMLST portal (https://pubmlst.org/). Phylotyping for *E. coli* was conducted using the ClermonTyping v20.3^22^ tool.

### Phylogenetic analysis

The assemblies were annotated with Bakta v1.9.2^23^, followed by pangenome analysis to identify core genes and generate a core gene alignment using Panaroo v1.3.3^24^ from which single nucleotide polymorphisms (SNPs) were extracted. The genetic relatedness among the *P*.

*mirabilis* isolates was assessed through phylogenetic analysis, for which a maximum likelihood tree was constructed using the extracted SNP data with the IQ-TREE v2.2.2.6^25^ with the ModelFinder Plus (MFP) option for identifying the optimal model for the data. Phylogenetic reconstruction also included information on invariant sites; branch supports were assessed with 1000 ultrafast bootstraps (UFBoot)^26^ approach implemented in IQ-TREE. The genetic relatedness for *E. coli* ST38 isolates was assessed using the same phylogenetic methods. The cophenetic distance was used to measure the evolutionary distance between the isolates connected by their most recent common ancestor (MRCA) on a phylogenetic tree. Then, these cophenetic distances were multiplied by the length of the core gene alignment to express the distance in terms of SNPs. More details of the phylogenetic analysis are available in the supplementary methods (see Supplementary Material).

### Mapping short-reads and genomic context

To assess the vehicles that potentially mobilize *bla*_OXA-23_ in *P. mirabilis* and *E. coli*, the genomic context was investigated by using Snippy v4.3.6^27^ to map sequence reads to a reference genome (complete VAC chromosome (GenBank: CP042907.1) as a reference for *P. mirabilis* and the complete chromosome of 1697008 (assembly: GCA_964341145) as a reference for *E. coli*) and reconstructing the consensus sequence for each isolate (details in supplementary methods [Supplementary Material]). The consensus sequence for each isolate was annotated with Bakta v1.9.2^23^ to identify insertion sequences (ISs) and AMR genes. The annotation of the transposable elements associated with *bla*_OXA-23_ in *P. mirabilis* was performed as described by Bonnin et al. 2020^5^. The complete chromosome sequence available for isolate 1697008 enabled the identification of the genomic context associated with *bla*_OXA-23_ in *E. coli*. This was achieved through manual annotation based on the genetic characteristics of transposable elements,^26^ including the presence of insertion sequences, terminal inverted repeat (IR) sequences, and target site duplications (TSD), also known as direct repeats (DR). Additionally, IslandViewer 4^28^ was used to identify genomic islands within the 1697008 genome. Finally, the genomic contexts of *bla*_OXA23_ were compared using Blastn v2.15.0^29^ and visualized along with their annotations using the genoPlotR v0.8.11^30^ R package.

## RESULTS AND DISCUSSION

### Clonal relatedness of OXA-23-producing *P. mirabilis* and *E. coli* isolates

The *bla*_OXA-23_ gene was identified in eight *P. mirabilis* and three *E. coli* (Table 1). A pangenome analysis of 64 OXA-23-producing *P. mirabilis* genomes resulted in a core gene alignment length of 2,513,546 bp (with 21,152 SNPs), representing 65% of the average total genome length of included sequences. Sixty-two of 64 isolates belonged to ST142, while the remaining two isolates were ST135 and ST185. There was substantial genetic diversity observed between isolates of different STs, with an average 13,791 SNPs between ST142 isolates and both ST135 and ST185, and 11,501 SNPs between the ST135 and ST185 isolates. All ST142 isolates formed a main cluster with some divergence between clades (Figure 1), with an average pairwise SNP difference of 102 between these isolates. This main ST142 lineage encompasses isolates derived from France, Belgium, Germany and the UK, as well as from diverse sources of human and animal origin.

**Table 1.**
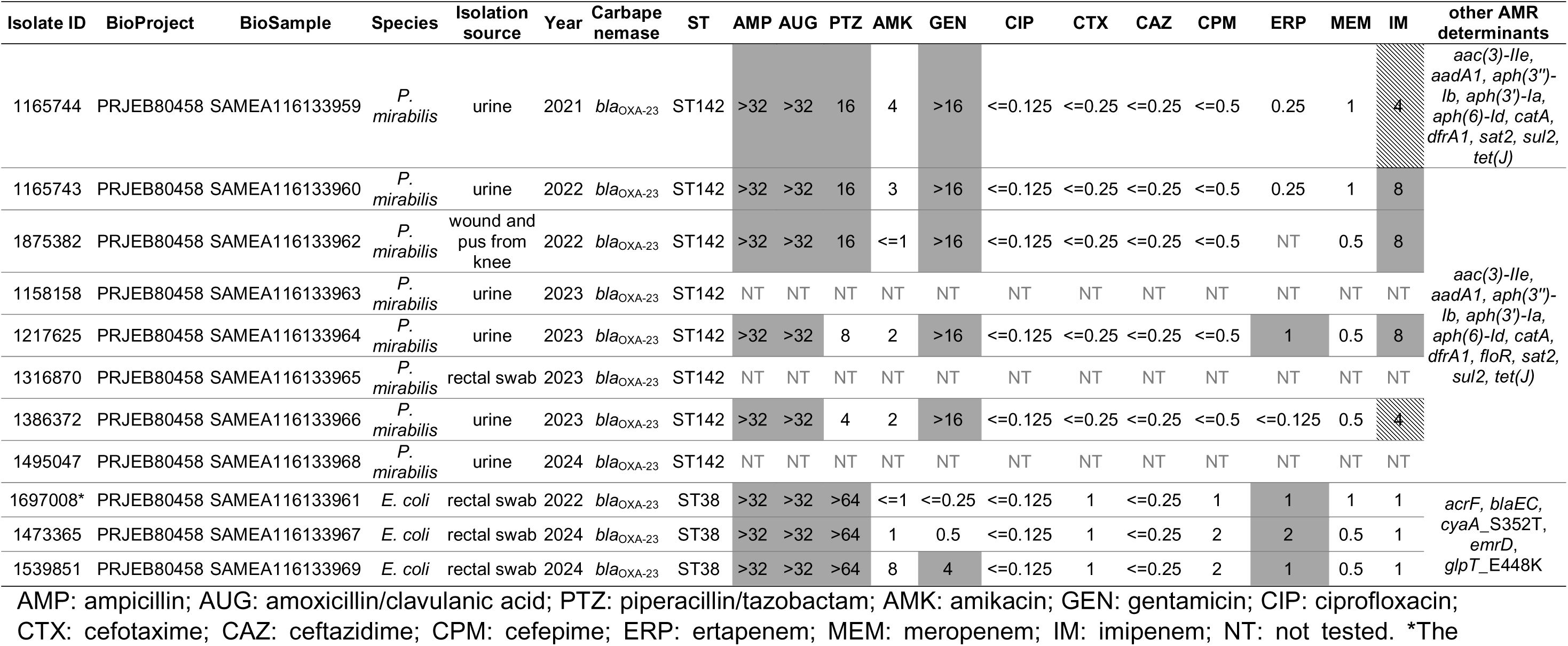
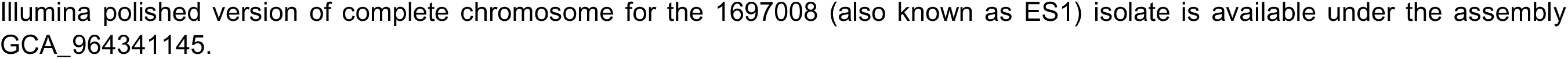
Accession numbers, metadata and antimicrobial susceptibility testing (AST) data for *P. mirabilis* (n=8) and *E. coli* (n=3) isolates obtained from human clinical samples in the UK. The table displays the minimum inhibitory concentration (MIC) values (in mg/L) for 12 antibiotics. MIC values were categorized according to the EUCAST^13^ guidelines using clinical breakpoints: 1) susceptible, indicated by the white box, 2) susceptible, increased exposure, indicated by the diagonal pattern, and 3) resistant, indicated by the grey box.

**Figure 1.**
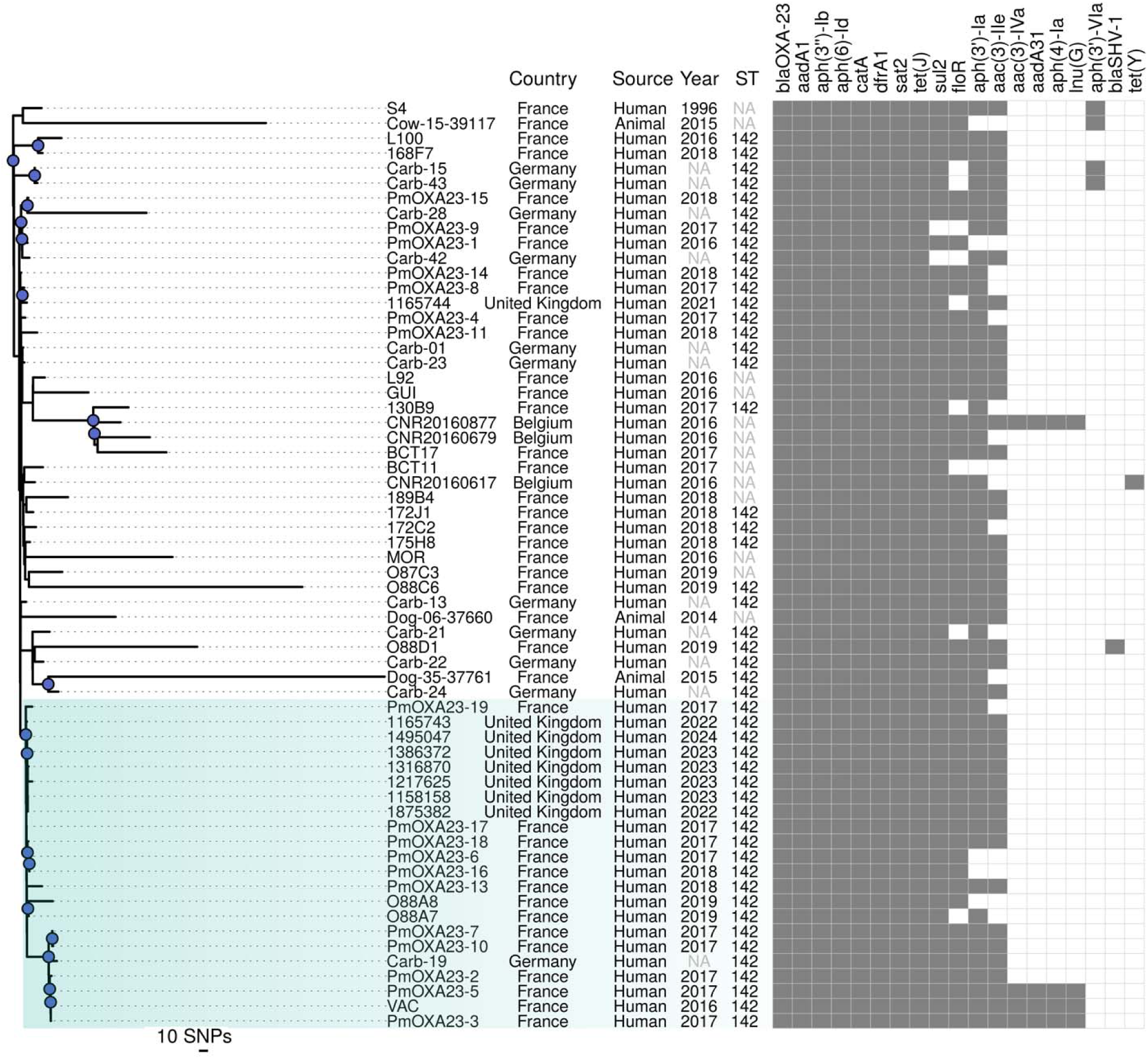
Core gene SNP-based rooted phylogeny of 62 *bla*_OXA-23_-positive *P. mirabilis* isolates. The phylogenetic relationship analysis included *bla*_OXA-23_-positive ST142 isolates from the UK (n=8) and isolates obtained from prior studies (n=54) conducted in various European countries. Next to the tree are represented the metadata such as country, source, year, ST and AMR determinants. “NA” label within year field, indicated that year-related information was not available, while “NA” in ST indicates that it was not feasible to establish the ST due do missing or incomplete MLST loci, this includes the absence or partial coverage (44%–86%) of *recA* in 11 genomes, the absence of *pyrC* in one genome or partial coverage (75%–58%) of *dnaJ* in two genomes, possibly attributable to fragmented contigs. Bootstrap support ≥95% (blue dots) indicates well supported clades, with the clade of interest highlighted in green.

Seven out of eight isolates from the UK were highly related, with a range of 0 – 9 pairwise SNPs, and grouped together in the tree. The remaining UK isolate exhibited slight divergence and was placed in a different cluster, with pairwise SNP distances ranging from 15 – 21 SNPs when compared with the group of seven highly similar isolates. In this study, a clade was defined as a group of isolates sharing a common ancestor and supported by a bootstrap value ≥95%. The ‘clade of interest’ was specifically designated to the clade containing the seven UK isolates and 15 international isolates, and showed a median pairwise SNP difference of 28 (IQR: 6 to 62) between the isolates. Within this clade of interest, when compared to the international isolates, the seven UK isolates were closely related to six French isolates (PmOXA23-19, PmOXA23-17, PmOXA23-18, PmOXA23-6, PmOXA23-16 and O88A7), with pairwise SNP distances ranging from 2 – 14 pairwise SNPs distance among them. This clade of interest includes the French VAC isolate, for which a complete genome is available. The French VAC isolate differed from the seven UK isolates by 29 – 34 SNPs (Figure 1). These relationships suggest that the isolates detected in the UK may have been introduced recently.

The first detection of *bla*_OXA-23_-positive *P. mirabilis* occurred in France during a study conducted between 1996-1999^4^. Subsequent sporadic detections were reported in clinical samples from 2016-2018, also in France. Notably, the affected patients had no history of international travel, indicating possible community acquisition of *bla*_OXA-23_-positive *P. mirabilis*^7^. Similarly, no travel history was reported for any of the patients from which UK isolates were recovered. *bla*_OXA-23_-positive *P. mirabilis* have subsequently been reported from twelve hospitals in France^6^, a routine screening sample in Finland in 2014^9^, clinical samples in Germany between 2013 and 2022^8^ and in Belgium and France, including animal samples, between 2014 and 2018^5^. More recently, *bla*_OXA-23_-positive *P. mirabilis* has been identified in clinical samples from Switzerland between 2017 and 2023^10^. Overall, these findings indicates that the expansion of the ST142 lineage has driven the spread of *bla*_OXA-23_-positive *P. mirabilis* in multiple countries and sources.

The core gene alignment of the UK *E. coli* isolates consisted of 3,353,881 bp (with 11,479 SNPs), representing 63% of the average total genome length of included sequences. All isolates belonged to ST38 and phylogroup D. The three *bla*_OXA-23_-positive isolates from geographically distinct regions of England (East Midlands, London, South East) were highly related, with pairwise distances between 5 – 9 SNPs. In comparison, when these three *bla*_OXA-23_-positive isolates were analyzed alongside available ST38 *bla*_OXA-23_-negative isolates, much larger pairwise genetic distances were observed [median (IQR) pairwise distance: 297 (216 to 723) SNPs]. To date, there have been only two previous reports of *bla*_OXA-23_ in *E. coli*: in ST4108 in Singapore (n=1)^11^ and ST471 in India (n=14)^12^; however, these studies did not generate WGS data for those isolates. To the best of our knowledge, our study is the first to report the presence of *bla*_OXA-23_ in *E. coli* ST38 in Europe.

### Antimicrobial susceptibility

In addition to *bla*_OXA-23_, the *P. mirabilis* ST142 isolates were found to harbor between 8 and 15 additional antimicrobial resistance genetic determinants (AMR genes; Figure 1; Table 1), which potentially confer resistance to eight different antimicrobial classes; nevertheless some of these AMR genes relate to intrinsic resistance (i.e. *tet(J)* to tetracycline and *cat* to phenicol)^5^.

All phenotypically tested *P. mirabilis* and *E. coli* isolates from this study were resistant to ampicillin (>32 mg/L) and amoxicillin/clavulanic acid (>32 mg/L). Most (6/8; 75%) isolates were also resistant to piperacillin/tazobactam (16 mg/L for *P. mirabilis* and >64 mg/L for *E. coli*). This aligns with the fact that OXA-23 enzyme can also hydrolyze penicillin class antibiotics^31^. All *P. mirabilis* isolates exhibited resistance to gentamicin, likely due to the presence of *aac(3)-IIe*. However, all isolates were susceptible to amikacin, ciprofloxacin, cefotaxime, ceftazidime and cefepime. This AST profile is expected as OXA-23 does not hydrolyze extended-spectrum cephalosporins^6^. The meropenem MICs of all isolates were within the susceptible range but above the EUCAST screening cut-off for investigation of suspected carbapenemase-producing Enterobacterales (>0.12 mg/L). However, all three *E. coli* and one *P. mirabilis* isolate were resistant to ertapenem, with MIC values ranging from 1 to 2 mg/L (Table 1). The observed low-level of carbapenem resistance agrees with the previous definition that OXA-23 is a weak carbapenem-hydrolyzing enzyme at least in *P. mirabilis*^5,6^, and that the bacterial host may impact on whether OXA enzymes are capable of hydrolyzing carbapenems^32^.

### Composite transposon harbouring *bla*_OXA-23_ in *P. mirabilis* ST142

The neighboring genes of *bla*_OXA-23_ were annotated in the flanking sequences, obtained using a short-read mapping approach with the French VAC *P. mirabilis* genome as a reference (Supplementary Figure S1). As previously reported in the French VAC genome^5^, *bla*_OXA-23_ is located within the transposon Tn*6704* (6,779 bp of length) with a gene configuration IS*Aba125*– IS*3*–IS*Aba14*–ATPase–*bla*_OXA-23_–IS*Aba1*–IS*4*, which is part of a larger composite transposon Tn*6703* (55,077 bp of length)^5^ (Figure 2A). The association between IS*Aba1* and *bla*_OXA-23_ has been well-documented in both *A. baumannii*^3^, and in *P. mirabilis*^5,7,9,10^ and IS*Aba1* is known to provide a promoter sequence that facilitates the expression of *bla*_OXA-23_^33^.

**Figure 2.**
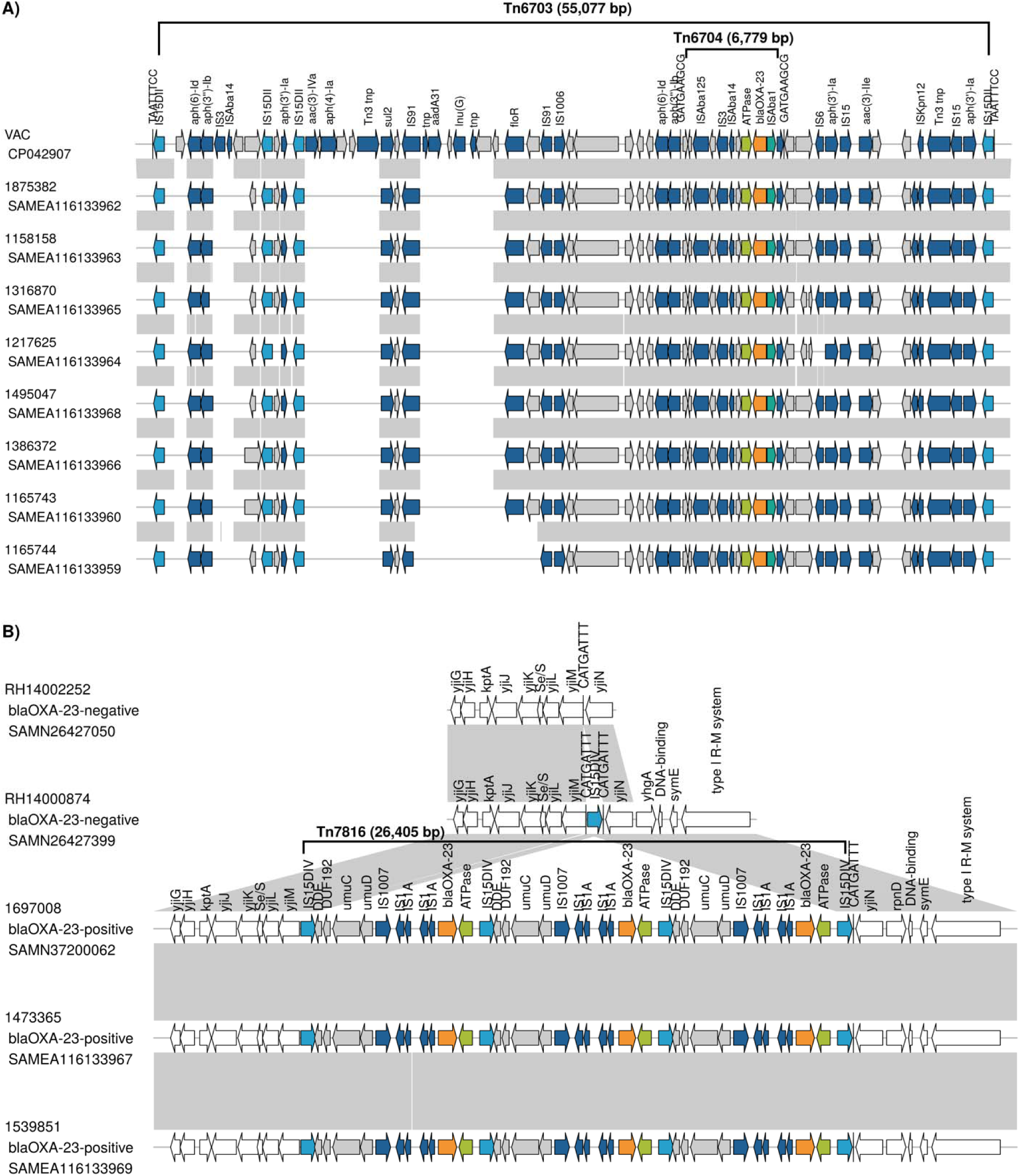
Genomic context associated with *bla*_OXA-23_ in the UK isolates. **A)** Composite transposon Tn*6703* present in *P. mirabilis* ST142 isolates from the UK (n=8) and France (with the VAC isolate as a reference) carrying AMR genes, and specifically Tn*6704*, which harbors *bla*_OXA-23_. Boundaries of Tn*6704* are indicated by the 9-bp target site duplication (TSD) GATGAAGCG and for Tn*6703* by 8-bp TSD TAATTTCC. **B)** A novel composite transposon Tn*7816* associated with *bla*_OXA-23_ in *bla*_OXA-23_-positive *E. coli* ST38 in UK isolates (n=3). The two *bla*_OXA-23_-negative isolates yielded contig-level sequence assemblies, where the operon *yjiGHJKLMN* and genetic configuration *yjiGHJKLM*–IS*15DIV*–*yjiN* were in a single contig, respectively. Genes are represented by arrows indicating the direction of transcription. *bla*_OXA-23_ is indicated by orange arrow, ATPase by light green arrow, IS*Aba1* by dark green arrow, IS*15DII* and IS*15DIV* by light blue arrows, other insertion sequences and additional AMR genes are indicated by dark blue arrows, other genes within the transposon by grey arrows, and other genes outside the transposon by white arrows. Grey areas between the linear plots represent nucleotide sequence identity (mostly >90%).

When comparing the VAC reference genome to the UK genomes, Tn*6703* was found to harbor six additional AMR genes in addition to *bla*_OXA-23_ in all genomes, conferring resistance to aminoglycosides (two copies each of *aph(6)-Id* [also known as *strB*] and *aph(3’’)-Ib* [also known as *strA*]; three copies of *aph(3’)-Ia* and one *aac(3)-IIe*), sulfonamides (*sul2*) and phenicol (*floR*, except one UK isolate). The VAC reference genome carried an additional four AMR genes [aminoglycosides: *aac(3)-IVa*, *aph(4)-Ia* and *aadA31*; lincosamides: (*lnu*(G)] that were absent from all UK isolates (Supplementary Figure S1). These findings align with previous observed variability in the gene content of the composite transposon Tn*6703* among the French^5^ and Swiss^10^ isolates.

### Novel composite transposon associated with *bla*_OXA-23_ in *E. coli* ST38

The genomic context of *bla*_OXA-23_ in *E. coli* ST38 from the UK was assessed using the 1697008 complete *E. coli* chromosome as a reference in the short-read mapping analysis. To further elucidate the transposon integration within the chromosome of the ST38 lineage, *bla*_OXA-23-_ negative *E. coli* ST38 genomes were also included in this study for comparative analysis (Supplementary Figure S2). This analysis revealed the integration of IS*15DIV* (which included left (IRL) and right (IRR) inverted terminal repeats sequence 14-bp GGCACTGTTGCAAA) within the chromosome of *bla*_OXA-23-_negative ST38, flanked by 8-bp TSD sequence CATGATTT. This integration disrupted the *yjiGHJKLMN* operon, resulting in the formation of a new genetic configuration, *yjiGHJKLM*–IS*15DIV*–*yjiN*, in these isolates (Figure 2B and Supplementary Figure S2). In the 1697008 isolate, along with the other two *bla*_OXA-23_-positive *E. coli*, *bla*_OXA-23_ was identified within a genomic island that corresponded to the boundaries of a novel transposon, which was named Tn*7816* according to the Transposon Registry database (https://transposon.lstmed.ac.uk/tn-registry). Tn*7816* is a composite transposon bordered by IS*15DIV* (which belongs to the IS*6* family transposase and is similar to IS*26*^34^) at both ends, flanked by 8-bp TSD sequence CATGATTT only at the right-end (Figure 2). This structure suggests a deletion of the left-end TSD sequence during transposition and rearrangement events. Tn*7816* carried three copies of *bla*_OXA-23_, each embedded within the specific genetic context: IS*15DIV*–DDE–DUF192–*umuC*–*umuD*–IS*1007*–IS*1*–IS*1A*–IS*1*–IS*1A*–*bla*_OXA-23_–ATPase (Figure 2). While this configuration differs from those previously reported in *A. baumannii*^3^ and *P. mirabilis*^5^, the presence of IS*1* linked with *bla*_OXA-23_ is consistent with a previous report of plasmid-borne *bla*_OXA-23_ in *E. coli*^11^. However, previous studies on *E. coli* ST4108 and ST471 were unable to characterize the detailed genetic context of plasmid-borne *bla*_OXA-23_ due to PCR assay limitations^11,12^. With all of this, we can hypothesize that *E. coli* ST38 might have acquired an OXA-23-encoding plasmid followed by integration of *bla*_OXA-23_ into the chromosome via mobile genetic elements, as reported here. A blastn search (https://blast.ncbi.nlm.nih.gov/Blast.cgi) against the NCBI Core nucleotide database (core_nt) identified *E. coli* ST457 isolate CE1628 (GenBank CP053852.1) from Australia (avian origin) with partial alignment with the transposon Tn*7816* sequence. However, in that genome the genomic context of *bla*_OXA-23_ (IS*15DIV* [inverse orientation]–DUF192–*umuC*–*umuD*–IS*1007*– IS*1*–IS*1A*–*bla*_OXA-23_–ATPase) and chromosomal location differed from the one investigated here. Specifically, the CE1628 isolate contains only a single copy of each *bla*_OXA-23_, IS*1* and IS*1A*.

Consequently, this study presents the first comprehensive characterization of this unique *bla*_OXA-23_ genetic context within *E. coli*.

In conclusion, our study underscores the significant role of transposon-carrying lineages in the emergence, dissemination and persistence of chromosomally-encoded *bla*_OXA-23_ in *P. mirabilis* and *E. coli*. Our findings provide key insights into three major aspects. First, the wide distribution of *bla*_OXA-23_ in *P. mirabilis* across multiple sources and countries, including the UK, France and Germany, suggesting international transmission of the *bla*_OXA-23_-positive *P. mirabilis* ST142 lineage. No epidemiological link could be established between the French, German and UK isolates, as they were collected in different years and none of the UK patients had a history of recent travel abroad. However, the high genetic relatedness between *P. mirabilis* ST142 isolates from the UK and France, along with the earlier observations within France, suggest a possible introduction of this lineage into the UK. Second, the association between the previously characterized transposon Tn*6704*, which includes IS*Aba1*, and the composite transposon Tn*6703*, which contains IS*15DII*, with *bla*_OXA-23_ in *P. mirabilis*, has been established. However, a notable variability in the gene content within Tn*6703* was observed, which may be attributed to the dynamic nature of transposable elements. Third, the emergence of *bla*_OXA-23_ in an *E. coli* ST38 lineage in the UK was facilitated by a novel composite transposon Tn*7816* characterized by the presence of IS*1*, IS*15DIV* and three copies of *bla*_OXA-23_. Together, these observations highlight the role of WGS in characterizing novel carbapenemase producers and associated mobile genetic elements, which may inform development of interventions to mitigate the spread of antimicrobial resistance.

Further studies are needed to monitor if *bla*_OXA-23_ becomes more widely established among Enterobacterales; however, surveillance is likely to be hampered by the lack of coverage of *bla*_OXA-23_ in commercial molecular assays^8^, which instead focus on the ‘big 5’ carbapenemase gene families. As per the UK Standards for Microbiological Investigations^35^, suspect Enterobacterales isolates exhibiting resistance to amoxicillin/clavulanate and/or reduced susceptibility to meropenem (MIC >0.12 mg/L) but negative for the ‘big 5’ carbapenemase gene families should be referred to the AMRHAI Reference Unit for further investigation.

## ABBREVIATIONS

AMR: antimicrobial resistance
AST: antimicrobial susceptibility testing
DR: direct repeats
ENA: European Nucleotide Archive
EUCAST: European Committee on Antimicrobial Susceptibility Testing
IQR: interquartile range
IR: inverted terminal repeats
MICs: minimum inhibitory concentrations
MLST: multi-locus sequence typing
PCR: polymerase chain reaction
SNPs: single nucleotide polymorphisms
ST: sequence type
TSD: target site duplication
WGS: whole genome sequencing
WHO: World Health Organization

## Data Availability

The sequencing data generated from this study were deposited under BioProject accession number PRJEB80458 in the ENA Project database. Accession numbers for all other sequence data used in this study are listed in Table S1.

## ACKNOWLEDGEMENTS

We gratefully acknowledge the use of the UKHSA core facilities HPC cluster servers. We thank hospital diagnostic laboratories for submitting the isolates reported here.

## AUTHOR CONTRIBUTIONS

KLH, KO, DW and NS secured funding to support the study. KLH, KO and DW designed the study. DM, AC and RP performed and oversaw microbiological testing. JFT carried out nanopore sequencing of isolate ES1/1697008. RZ performed the data analysis. RZ and KLH wrote the manuscript, and all authors edited and approved the final draft of the manuscript.

## ETHICAL STATEMENT

Not applicable.

