## Supplementary data for "Dissemination of OXA-23 carbapenemase-producing *Proteus mirabilis* and *Escherichia coli* is driven by transposon-carrying lineages in the UK"

### **SUPPLEMENTARY METHODS**

#### **Whole genome sequence data**

This study generated Illumina short-reads for *bla*_OXA-23_-positive *P. mirabilis* (n=8) and *E. coli* (n=3) isolates from the UK. In regards the publicly available genomes, the fastq files for French *bla*_OXA-23_-posititve *P. mirabilis*^1^ (Bioproject PRJNA490489), and UK *bla*_OXA-23_-negative *E. coli* ST38^2^ (Bioproject PRJNA812750) isolates were accessible and downloaded from the European Nucleotide Archive (ENA). While contigs files were available for *bla*_OXA-23_-positive *P. mirabilis* from France and Belgium^3^ (Bioproject PRJNA521327), France^4^ (Bioproject PRJNA780406) and Germany^5^ (Bioproject PRJNA915754). These contigs files were downloaded from NCBI/GenBank.

A complete chromosome sequence was publicly available for a *bla*_OXA-23_-positive *P. mirabilis* VAC isolate (Genbank CP042907.1) from France^3^ and a *bla*_OXA-23_-positive *E. coli* 1697008 (also known as ES1) isolate (Genbank CP133856.1) from the UK. To obtain highly accuracy of 1697008 (ES1) long-read-only assembly, a hybrid polishing approach utilizing Illumina short-reads and the Pilon v1.24^6^ tool was employed. A single round of hybrid polishing was enough to identify and correct 71 mismatches (31 SNPs and 40 indels) in the assembly, resulting in an Illumina polished version of the complete chromosome for the 1697008 (ES1) isolate. The Illumina polished chromosome sequence was subsequently deposited into the ENA under the assembly accession number GCA_964341145.

#### **Pangenome and phylogenetic analysis**

Draft genomes and complete chromosomes were annotated using Bakta v1.9.2^7^. The pangenome was independently analysed for each *P. mirabilis* and *E. coli* dataset using Panaroo v1.3.3^8^ with --clean-mode strict parameter for conservative approach, which removes potential contamination and incorrect annotation. The identification of core genes was based on 98% sequence similarity and their presence in 95% of isolates. Subsequently, this resulted on a filtered core gene alignment, in which the problematic regions are already removed if they exceed the Block Mapping and Gathering with Entropy (BMGE) filter^8^, which is recommended for building core gene phylogenies. To assess the phylogenetic relatedness among the isolates, single nucleotide polymorphisms (SNPs) were extracted from the filtered core gene alignment with the tool snp-sites v2.5.2^9^. Then, these SNPs were used to construct a maximum likelihood tree in IQ-TREE v2.2.2.6^10^. The ModelFinder Plus (MFP) option was employed to identify the best-fitting model for the data, and the selected model was then used to reconstruct the tree. In addition to the -m MFP parameter, the following options were included: -mrate E,I,R -cmin 2 -cmax 10, which define the type of rate heterogeneity and the range for the number of categories in the FreeRate model. Information on constant sites and 1000 ultrafast bootstraps (UFBoot) were also included in the analysis. The results of the MFP assessment showed that the best substitution model for both *P. mirabilis* and *E. coli* was GTR (General time reversible model). For *P. mirabilis*, the rate heterogeneity was modeled using +I, which allows for a proportion of invariable sites, and for *E. coli* it was modeled with +R7 using the FreeRate approach. The phylogenetic trees were constructed using these models. The *P. mirabilis* tree was rooted using the 160A10 isolate (ST185) as an outgroup, while the *E. coli* tree was midpoint rooting as no outgroup was used. The trees were plotted alongside with the metadata (country, source, year, ST and presence/absence of AMR genes, and other relevant data) using ggtree v3.12.0^11^ R^12^ package.

The genetic distance among *bla*_OXA-23_-positive *P. mirabilis* isolates was assessed by calculating the cophenetic distance by using the cophenetic() function available in the stats package from R^12^. The cophenetic distance measures the evolutionary distance between pair of taxa (or 'tips') on a phylogenetic tree. It is calculated by measuring the length of the branch connecting their most recent common ancestor (MRCA)^13^. Then, the cophenetic distance is multiplied by the length of the core gene alignment to express the distance in SNPs. Similarly, the genetic distance among the *E. coli* isolates were assessed as described above.

#### **Consensus sequence obtained by short-read mapping**

The short-reads were mapped to a reference genome to obtain the consensus sequence for each isolate. A complete chromosome of the VAC isolate (Genbank CP042907.1) from France^3^ served as a reference for *bla*_OXA-23_-positive *P. mirabilis* isolates (UK n=8 and France n=18), and the complete chromosome of 1697008 (ES1) isolate (Assembly GCA_964341145 ) from the UK was used as a reference for *E. coli* ST38 isolates (*bla*_OXA-23_-positive n=3 and *bla*_OXA-23_-negative n=32; all isolates from the UK). Short-read data were mapped against the reference using Snippy v4.3.6^14^ including the settings --mincov 5 --mapqual 0, which allowed for a minimum depth coverage of 5x and retrieval of sequences from multi-mapping regions. This methodology enabled the acquisition of a consensus sequence for each isolate individually. These individual sequences were aligned using Snippy-core and generating thus the core genome alignment. The genomic neighbourhood around *bla*_OXA-23_ gene were identified and visually presented as a heatmap adjacent to the phylogenetic tree. The consensus sequences were annotated with Bakta v1.9.2^7^ and used for the analysis of the genomic context of *bla*_OXA-23_ described in the main text.

### **SUPPLEMENTARY FIGURES**

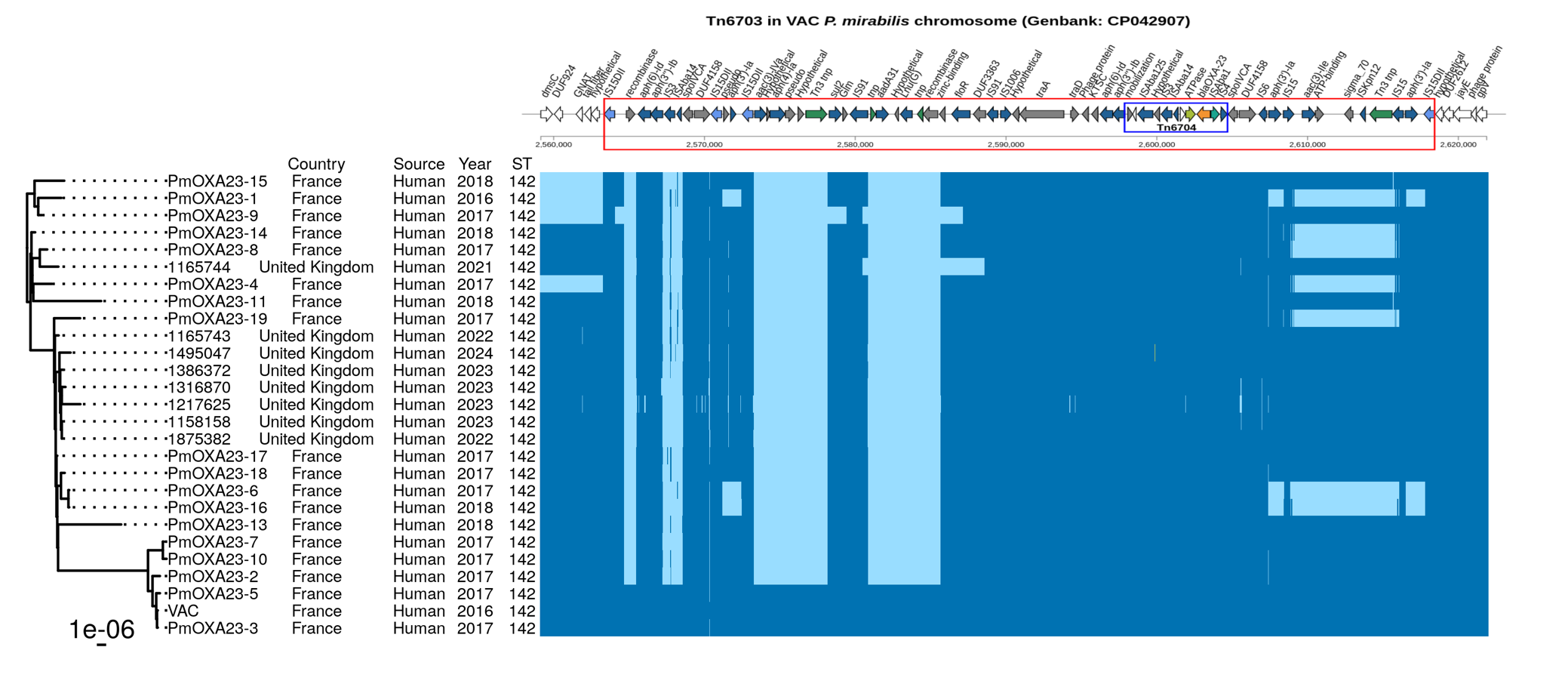

**Supplementary Figure S1. Mapping short-reads for ST142 isolates against the reference VAC *bla*_OXA-23_-positive *P. mirabilis* genome.** Core gene SNP-based phylogeny of 27 ST142 *bla*_OXA-23_-positive *P. mirabilis* genomes. The UK isolates were obtained from our study, while the French isolates were from Potron et al. 2019^1^. The accompanying metadata alongside the phylogenetic tree contains information on country, source, year, and sequence type (ST). The heatmap represents the genomic neighbourhood around the *bla*_OXA-23_ obtained through the short-read mapping approach, which includes the composite transposon Tn*6703* and five genes upstream and downstream. In the heatmap, dark blue represents an identical sequence to the reference, while light blue indicates an absence of the sequence. Annotations at the top of the heatmap are represented by coloured arrows, with *bla*_OXA-23_ represented in orange, ATPase in light green, IS*Aba1* in dark green, IS*15DII* in light blue, other insertion sequences and additional antimicrobial resistance (AMR) genes in dark blue, other genes in grey, and the five genes located upstream and downstream of Tn*6703* in white. The transposons Tn*6703* and Tn*6704* are visually indicated by red and blue rectangles, respectively.

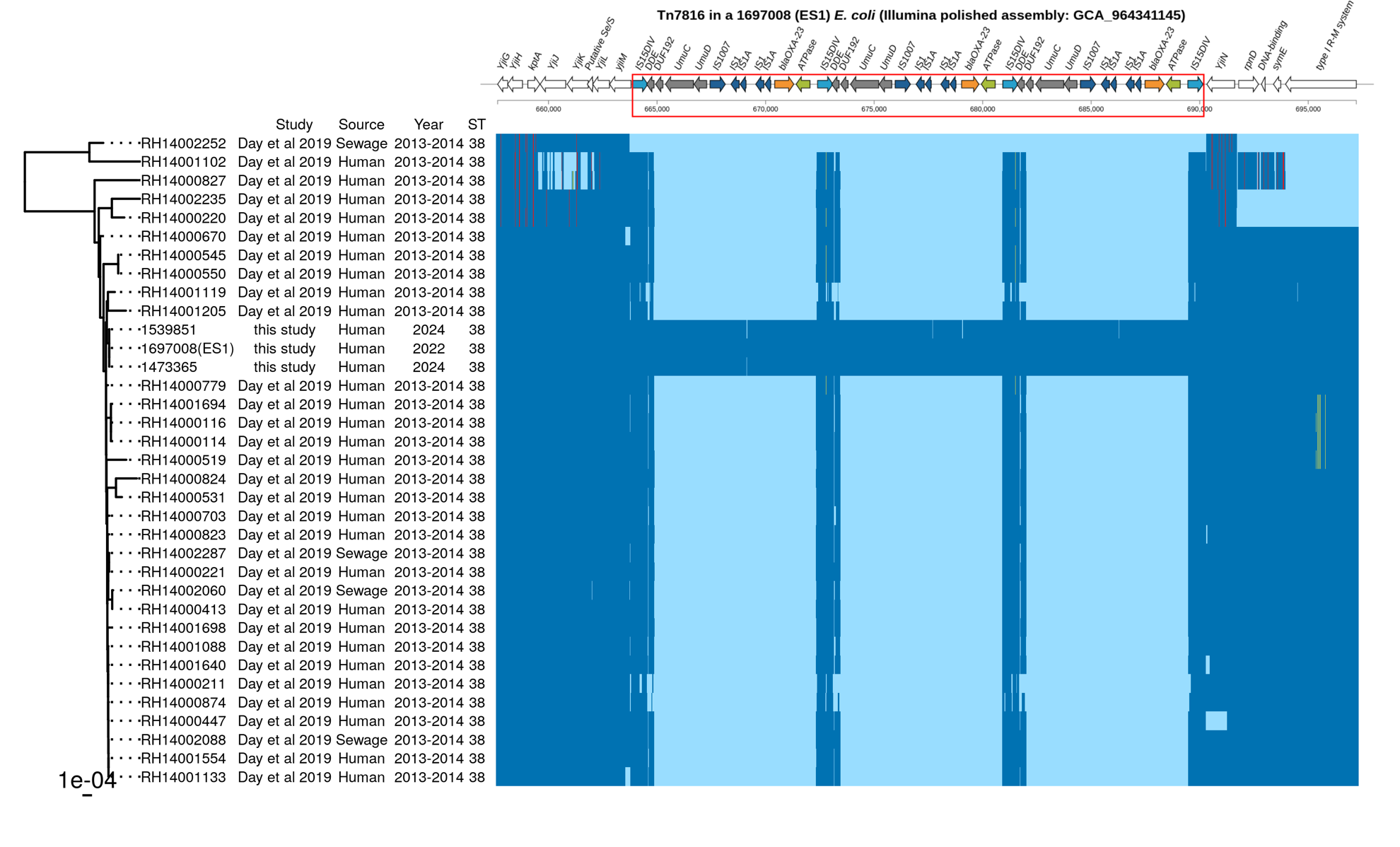

**Supplementary Figure S2. Mapping short-reads for ST38 isolates against the reference 1697008 (also known as ES1) *bla*_OXA-23_-positive *E. coli* genome**. Core gene SNP-based phylogeny of 35 ST38 *E. coli* genomes from the UK. The *bla*_OXA-23_-positive isolates (n=3) were from our study, while the *bla*_OXA-23_-negative isolates (n=32) were from Day et al. 2019^2^. The accompanying metadata alongside the phylogenetic tree contains information on the study, source, year, and sequence type (ST). The heatmap represents the genomic neighbourhood around *bla*_OXA-23_ obtained through the short-read mapping approach, which includes the novel composite transposon Tn*7816* and eight genes upstream and five downstream. In the heatmap, dark blue represents an identical sequence to the reference, while light blue indicates an absence of the sequence. Annotations at the top of the heatmap are represented by coloured arrows, with *bla*_OXA-23_ represented in orange, ATPase in light green, IS*15DIV* in light blue, other insertion sequences in dark blue, other genes in grey, and the eight genes located upstream and five genes downstream of Tn*7816* in white. The novel composite transposon Tn*7816* is visually indicated by red rectangle.

### **SUPPLEMENTARY TABLE**

**Supplementary Table S1. List of *P. mirabilis* (n=56) and *E. coli* (n=32) genomes from previous studies, including accession numbers and metadata.** All *P. mirabilis* harbour *bla*_OXA-23_, while none of the *E. coli* isolates do.

| **Isolate ID** | **Species** | **BioProject** | **BioSample** | **Assembly** | **SRA** | **Source** | **Country** | **Year** | **ST** | **Study** |
| --- | --- | --- | --- | --- | --- | --- | --- | --- | --- | --- |
| 130B9 | *P. mirabilis* | PRJNA521327 | SAMN10883386 | GCA_004570075.1 |  | Human | France | 2017 | 142 | Bonnin et al. 2020^3^ |
| 160A10 | *P. mirabilis* | PRJNA521327 | SAMN10883387 | GCA_004570785.1 |  | Human | France | 2018 | 185 |  |
| 168F7 | *P. mirabilis* | PRJNA521327 | SAMN10883388 | GCA_004570225.1 |  | Human | France | 2018 | 142 |  |
| 172C2 | *P. mirabilis* | PRJNA521327 | SAMN10883389 | GCA_004570215.1 |  | Human | France | 2018 | 142 |  |
| 172J1 | *P. mirabilis* | PRJNA521327 | SAMN10883390 | GCA_004570745.1 |  | Human | France | 2018 | 142 |  |
| 175H8 | *P. mirabilis* | PRJNA521327 | SAMN10883391 | GCA_004570715.1 |  | Human | France | 2018 | 142 |  |
| 189B4 | *P. mirabilis* | PRJNA521327 | SAMN10883393 | GCA_004570665.1 |  | Human | France | 2018 | NA |  |
| BCT11 | *P. mirabilis* | PRJNA521327 | SAMN10883407 | GCA_004569245.1 |  | Human | France | 2017 | NA |  |
| BCT17 | *P. mirabilis* | PRJNA521327 | SAMN10883408 | GCA_004569205.1 |  | Human | France | 2017 | NA |  |
| CNR20160617 | *P. mirabilis* | PRJNA521327 | SAMN10883403 | GCA_004570145.1 |  | Human | Belgium | 2016 | NA |  |
| CNR20160679 | *P. mirabilis* | PRJNA521327 | SAMN10883401 | GCA_004570175.1 |  | Human | Belgium | 2016 | NA |  |
| CNR20160877 | *P. mirabilis* | PRJNA521327 | SAMN10883402 | GCA_004570235.1 |  | Human | Belgium | 2016 | NA |  |
| Cow-15-39117 | *P. mirabilis* | PRJNA521327 | SAMN10883396 | GCA_004570065.1 |  | Animal | France | 2015 | NA |  |
| Dog-06-37660 | *P. mirabilis* | PRJNA521327 | SAMN10883397 | GCA_004570055.1 |  | Animal | France | 2014 | NA |  |
| Dog-35-37761 | *P. mirabilis* | PRJNA521327 | SAMN10883398 | GCA_004570195.1 |  | Animal | France | 2015 | 142 |  |
| GUI | *P. mirabilis* | PRJNA521327 | SAMN10883404 | GCA_004570115.1 |  | Human | France | 2016 | NA |  |
| L100 | *P. mirabilis* | PRJNA521327 | SAMN10883399 | GCA_004570645.1 |  | Human | France | 2016 | 142 |  |
| L92 | *P. mirabilis* | PRJNA521327 | SAMN10883400 | GCA_004570275.1 |  | Human | France | 2016 | NA |  |
| MOR | *P. mirabilis* | PRJNA521327 | SAMN10883406 | GCA_004569255.1 |  | Human | France | 2016 | NA |  |
| S4 | *P. mirabilis* | PRJNA521327 | SAMN10883394 | GCA_004570775.1 |  | Human | France | 1996 | NA |  |
| VAC* | *P. mirabilis* | PRJNA521327 | SAMN12566310 | GCA_008041895.1 |  | Human | France | 2016 | 142 |  |
| Carb-01 | *P. mirabilis* | PRJNA915754 | SAMN32405799 | GCA_030336315.1 |  | Human | Germany | NA | 142 | Hamprecht et al. 2023^5^ |
| Carb-13 | *P. mirabilis* | PRJNA915754 | SAMN32405811 | GCA_030336105.1 |  | Human | Germany | NA | 142 |  |
| Carb-15 | *P. mirabilis* | PRJNA915754 | SAMN32405813 | GCA_030336145.1 |  | Human | Germany | NA | 142 |  |
| Carb-17 | *P. mirabilis* | PRJNA915754 | SAMN32405815 | GCA_030336065.1 |  | Human | Germany | NA | 135 |  |
| Carb-19 | *P. mirabilis* | PRJNA915754 | SAMN32405817 | GCA_030336005.1 |  | Human | Germany | NA | 142 |  |
| Carb-21 | *P. mirabilis* | PRJNA915754 | SAMN32405819 | GCA_030336025.1 |  | Human | Germany | NA | 142 |  |
| Carb-22 | *P. mirabilis* | PRJNA915754 | SAMN32405820 | GCA_030335965.1 |  | Human | Germany | NA | 142 |  |
| Carb-23 | *P. mirabilis* | PRJNA915754 | SAMN32405821 | GCA_030335945.1 |  | Human | Germany | NA | 142 |  |
| Carb-24 | *P. mirabilis* | PRJNA915754 | SAMN32405822 | GCA_030335905.1 |  | Human | Germany | NA | 142 |  |
| Carb-28 | *P. mirabilis* | PRJNA915754 | SAMN32405826 | GCA_030335865.1 |  | Human | Germany | NA | 142 |  |
| Carb-42 | *P. mirabilis* | PRJNA915754 | SAMN32405840 | GCA_030335585.1 |  | Human | Germany | NA | 142 |  |
| Carb-43 | *P. mirabilis* | PRJNA915754 | SAMN32405841 | GCA_030335605.1 |  | Human | Germany | NA | 142 |  |
| O87C3 | *P. mirabilis* | PRJNA780406 | SAMN23139460 | GCA_021095805.1 |  | Human | France | 2019 | NA | Lombes et al. 2022^4^ |
| O88A7 | *P. mirabilis* | PRJNA780406 | SAMN23139465 | GCA_021095725.1 |  | Human | France | 2019 | 142 |  |
| O88A8 | *P. mirabilis* | PRJNA780406 | SAMN23139466 | GCA_021095705.1 |  | Human | France | 2019 | 142 |  |
| O88C6 | *P. mirabilis* | PRJNA780406 | SAMN23139468 | GCA_021095635.1 |  | Human | France | 2019 | 142 |  |
| O88D1 | *P. mirabilis* | PRJNA780406 | SAMN23139469 | GCA_021095615.1 |  | Human | France | 2019 | 142 |  |
| PmOXA23-1 | *P. mirabilis* | PRJNA490489 | SAMN10038483 |  | SRS3767930 | Human | France | 2016 | 142 | Potron et al. 2019^1^ |
| PmOXA23-10 | *P. mirabilis* | PRJNA490489 | SAMN10038492 |  | SRS3767937 | Human | France | 2017 | 142 |  |
| PmOXA23-11 | *P. mirabilis* | PRJNA490489 | SAMN10038493 |  | SRS3767920 | Human | France | 2018 | 142 |  |
| PmOXA23-13 | *P. mirabilis* | PRJNA490489 | SAMN10038495 |  | SRS3767923 | Human | France | 2018 | 142 |  |
| PmOXA23-14 | *P. mirabilis* | PRJNA490489 | SAMN10038496 |  | SRS3767922 | Human | France | 2018 | 142 |  |
| PmOXA23-15 | *P. mirabilis* | PRJNA490489 | SAMN10038497 |  | SRS3767926 | Human | France | 2018 | 142 |  |
| PmOXA23-16 | *P. mirabilis* | PRJNA490489 | SAMN10038498 |  | SRS3767924 | Human | France | 2018 | 142 |  |
| PmOXA23-17 | *P. mirabilis* | PRJNA490489 | SAMN10038499 |  | SRS3767925 | Human | France | 2017 | 142 |  |
| PmOXA23-18 | *P. mirabilis* | PRJNA490489 | SAMN10038500 |  | SRS3767927 | Human | France | 2017 | 142 |  |
| PmOXA23-19 | *P. mirabilis* | PRJNA490489 | SAMN10038501 |  | SRS3767919 | Human | France | 2017 | 142 |  |
| PmOXA23-2 | *P. mirabilis* | PRJNA490489 | SAMN10038484 |  | SRS3767928 | Human | France | 2017 | 142 |  |
| PmOXA23-3 | *P. mirabilis* | PRJNA490489 | SAMN10038485 |  | SRS3767931 | Human | France | 2017 | 142 |  |
| PmOXA23-4 | *P. mirabilis* | PRJNA490489 | SAMN10038486 |  | SRS3767929 | Human | France | 2017 | 142 |  |
| PmOXA23-5 | *P. mirabilis* | PRJNA490489 | SAMN10038487 |  | SRS3767933 | Human | France | 2017 | 142 |  |
| PmOXA23-6 | *P. mirabilis* | PRJNA490489 | SAMN10038488 |  | SRS3767932 | Human | France | 2017 | 142 |  |
| PmOXA23-7 | *P. mirabilis* | PRJNA490489 | SAMN10038489 |  | SRS3767935 | Human | France | 2017 | 142 |  |
| PmOXA23-8 | *P. mirabilis* | PRJNA490489 | SAMN10038490 |  | SRS3767934 | Human | France | 2017 | 142 |  |
| PmOXA23-9 | *P. mirabilis* | PRJNA490489 | SAMN10038491 |  | SRS3767936 | Human | France | 2017 | 142 |  |
| RH14000114 | *E. coli* | PRJNA812750 | SAMN26427511 |  | SRS12339957 | Human | UK | 2013-2014 | 38 | Day et al. 2019^2^ |
| RH14000116 | *E. coli* | PRJNA812750 | SAMN26427512 |  | SRS12339958 | Human | UK | 2013-2014 | 38 |  |
| RH14000211 | *E. coli* | PRJNA812750 | SAMN26426995 |  | SRS12340023 | Human | UK | 2013-2014 | 38 |  |
| RH14000220 | *E. coli* | PRJNA812750 | SAMN26427015 |  | SRS12340046 | Human | UK | 2013-2014 | 38 |  |
| RH14000221 | *E. coli* | PRJNA812750 | SAMN26426872 |  | SRS12339696 | Human | UK | 2013-2014 | 38 |  |
| RH14000413 | *E. coli* | PRJNA812750 | SAMN26426928 |  | SRS12340221 | Human | UK | 2013-2014 | 38 |  |
| RH14000447 | *E. coli* | PRJNA812750 | SAMN26427304 |  | SRS12340175 | Human | UK | 2013-2014 | 38 |  |
| RH14000519 | *E. coli* | PRJNA812750 | SAMN26427770 |  | SRS12340084 | Human | UK | 2013-2014 | 38 |  |
| RH14000531 | *E. coli* | PRJNA812750 | SAMN26427345 |  | SRS12339839 | Human | UK | 2013-2014 | 38 |  |
| RH14000545 | *E. coli* | PRJNA812750 | SAMN26427625 |  | SRS12339532 | Human | UK | 2013-2014 | 38 |  |
| RH14000550 | *E. coli* | PRJNA812750 | SAMN26426921 |  | SRS12339535 | Human | UK | 2013-2014 | 38 |  |
| RH14000670 | *E. coli* | PRJNA812750 | SAMN26426945 |  | SRS12339374 | Human | UK | 2013-2014 | 38 |  |
| RH14000703 | *E. coli* | PRJNA812750 | SAMN26426906 |  | SRS12339495 | Human | UK | 2013-2014 | 38 |  |
| RH14000779 | *E. coli* | PRJNA812750 | SAMN26427420 |  | SRS12339664 | Human | UK | 2013-2014 | 38 |  |
| RH14000823 | *E. coli* | PRJNA812750 | SAMN26427788 |  | SRS12340104 | Human | UK | 2013-2014 | 38 |  |
| RH14000824 | *E. coli* | PRJNA812750 | SAMN26427783 |  | SRS12340099 | Human | UK | 2013-2014 | 38 |  |
| RH14000827 | *E. coli* | PRJNA812750 | SAMN26427784 |  | SRS12340100 | Human | UK | 2013-2014 | 38 |  |
| RH14000874 | *E. coli* | PRJNA812750 | SAMN26427399 |  | SRS12339643 | Human | UK | 2013-2014 | 38 |  |
| RH14001088 | *E. coli* | PRJNA812750 | SAMN26427580 |  | SRS12340001 | Human | UK | 2013-2014 | 38 |  |
| RH14001102 | *E. coli* | PRJNA812750 | SAMN26427025 |  | SRS12340284 | Human | UK | 2013-2014 | 38 |  |
| RH14001119 | *E. coli* | PRJNA812750 | SAMN26427734 |  | SRS12339393 | Human | UK | 2013-2014 | 38 |  |
| RH14001133 | *E. coli* | PRJNA812750 | SAMN26427303 |  | SRS12340174 | Human | UK | 2013-2014 | 38 |  |
| RH14001205 | *E. coli* | PRJNA812750 | SAMN26426978 |  | SRS12339686 | Human | UK | 2013-2014 | 38 |  |
| RH14001554 | *E. coli* | PRJNA812750 | SAMN26427341 |  | SRS12339834 | Human | UK | 2013-2014 | 38 |  |
| RH14001640 | *E. coli* | PRJNA812750 | SAMN26427586 |  | SRS12340007 | Human | UK | 2013-2014 | 38 |  |
| RH14001694 | *E. coli* | PRJNA812750 | SAMN26427359 |  | SRS12339853 | Human | UK | 2013-2014 | 38 |  |
| RH14001698 | *E. coli* | PRJNA812750 | SAMN26427055 |  | SRS12339575 | Human | UK | 2013-2014 | 38 |  |
| RH14002060 | *E. coli* | PRJNA812750 | SAMN26427331 |  | SRS12339498 | Sewage | UK | 2013-2014 | 38 |  |
| RH14002088 | *E. coli* | PRJNA812750 | SAMN26427561 |  | SRS12340237 | Sewage | UK | 2013-2014 | 38 |  |
| RH14002235 | *E. coli* | PRJNA812750 | SAMN26427380 |  | SRS12339908 | Human | UK | 2013-2014 | 38 |  |
| RH14002252 | *E. coli* | PRJNA812750 | SAMN26427050 |  | SRS12339569 | Sewage | UK | 2013-2014 | 38 |  |
| RH14002287 | *E. coli* | PRJNA812750 | SAMN26427711 |  | SRS12339367 | Sewage | UK | 2013-2014 | 38 |  |

*The chromosome sequence for VAC isolate is available under the GenBank CP042907.1.

The “NA” label in the Year column indicates that the corresponding data was not available. Particularly, the German samples were collected between 2013 to 2022, however the specific collection year for each sample was not provided^5^. Similarly, the UK samples from Day et al. 2019^2^ study were collected between 2013 and 2014, but information regarding the individual collection year for each sample was not available.

The "NA" in the ST column indicates that the sequence type could not be assigned due to the absence of necessary MLST loci, likely a result of fragmented genome assemblies.
